# Substance abuse and the risk of severe COVID-19: Mendelian randomization confirms the causal role of opioids but hints a negative causal effect for cannabinoids

**DOI:** 10.1101/2022.05.06.22274584

**Authors:** M.Reza Jabalameli, Zhengdong D Zhang

## Abstract

Since the start of the COVID-19 global pandemic, our understanding of the underlying disease mechanism and factors associated with the disease severity has dramatically increased. A recent report investigated the relationship between substance use disorders (SUD) and the risk of severe COVID-19 in the United States and concluded that the risk of hospitalization and death due to COVID-19 is directly correlated with substance abuse, including opioid use disorder (OUD) and cannabis use disorder (CUD). While we found this analysis fascinating, we believe this observation may be biased due to comorbidities (such as hypertension, diabetes, and cardiovascular disease) confounding the direct impact of SUD on severe COVID-19 illness. To objectively answer this question, we sought to investigate the causal relationship between substance abuse and medication-taking history (as a proxy trait for comorbidities) with the risk of COVID-19 adverse outcomes. Our Mendelian randomization analysis confirms the causal relationship between SUD and severe COVID-19 illness but hints at a negative causal effect for cannabinoids. Given that a great deal of COVID-19 mortality is attributed to disturbed immune regulation, the possible modulatory impact of cannabinoids in alleviating cytokine storms merits further investigation.

## Main

Since the start of the COVID-19 global pandemic, our understanding of the underlying disease mechanism and factors associated with the disease severity has dramatically increased. In a timely study, *Wang et al*.[1] investigated the relationship between substance use disorders (SUD) and the risk of severe COVID-19 in the United States and concluded that the risk of hospitalization and death due to COVID-19 is directly correlated with substance abuse, particularly with opioid use disorder. While we found this analysis fascinating, we believe this observation may be biased due to comorbidities (such as hypertension, diabetes, cardiovascular disease (CVD), etc.) confounding the direct impact of SUD on severe COVID-19 illness.

To objectively test whether drug abuse is causally related to an increased risk of COVID-19 adverse outcomes, we carried out Mendelian randomization (MR) analysis using available high powered GWAS analysis of substance abuse (Cannabinoids[2], Opioids[3], Alcohol), medication-taking history[4] (as a proxy trait for comorbidities such as CVD) and the risk of Covid-19 hospitalization and respiratory failure5 (COVID-19 Host Genetic Initiative- release 4) (See **Supplementary Note**). MR uses exposure-associated genetic variants as instrumental variables to investigate the causal relationship between the exposure and outcome[5]. Since genetic variants are randomly segregated at conception, MR resembles randomized controlled trials but is more robust to confounding than observational studies (See **Supplementary Note**).

The motivation behind our analysis is to circumvent the confounding imparted by unmeasured comorbidities in the investigation of the causal relationship between SUD and COVID-19 adverse outcomes. The major limitation of *Wang et al*. study is that they could not control for comorbidities due to the size limitation. Thus, although they reframed their null hypothesis to test whether SUD associated comorbidities contribute to patients’ risk of COVID-19 adverse outcomes, the present observational analysis cannot assess the relationship between SUD and COVID-19 illness; it is not clear whether the observed relationship is due to the higher prevalence of comorbidities among cases (compared to controls), or substance abuse has a real causal effect on COVID-19 severe illness.

To answer this question, we investigated the causality between four SUD classes – i.e., opium use disorder (OUD), cannabis use disorder (CUD), alcohol use disorder (AUD) – and COVID-19 hospitalization and severe respiratory outcomes. We also investigated the causal relationship of six medication categories (including opioids) with COVID-19 outcomes. We carried out the MR analysis using four different methods: LCV[5], Egger regression[6], inverse-variance weighted (IVW) regression8, and weighted median[7]. MR analysis in this context enabled us to investigate the true nature of causality while controlling for comorbidities. Presented in **Table 1** are exposure traits with a significant causal relationship with severe COVID-19 indicated by at least one MR method after Bonferroni correction.

**Table 1.**
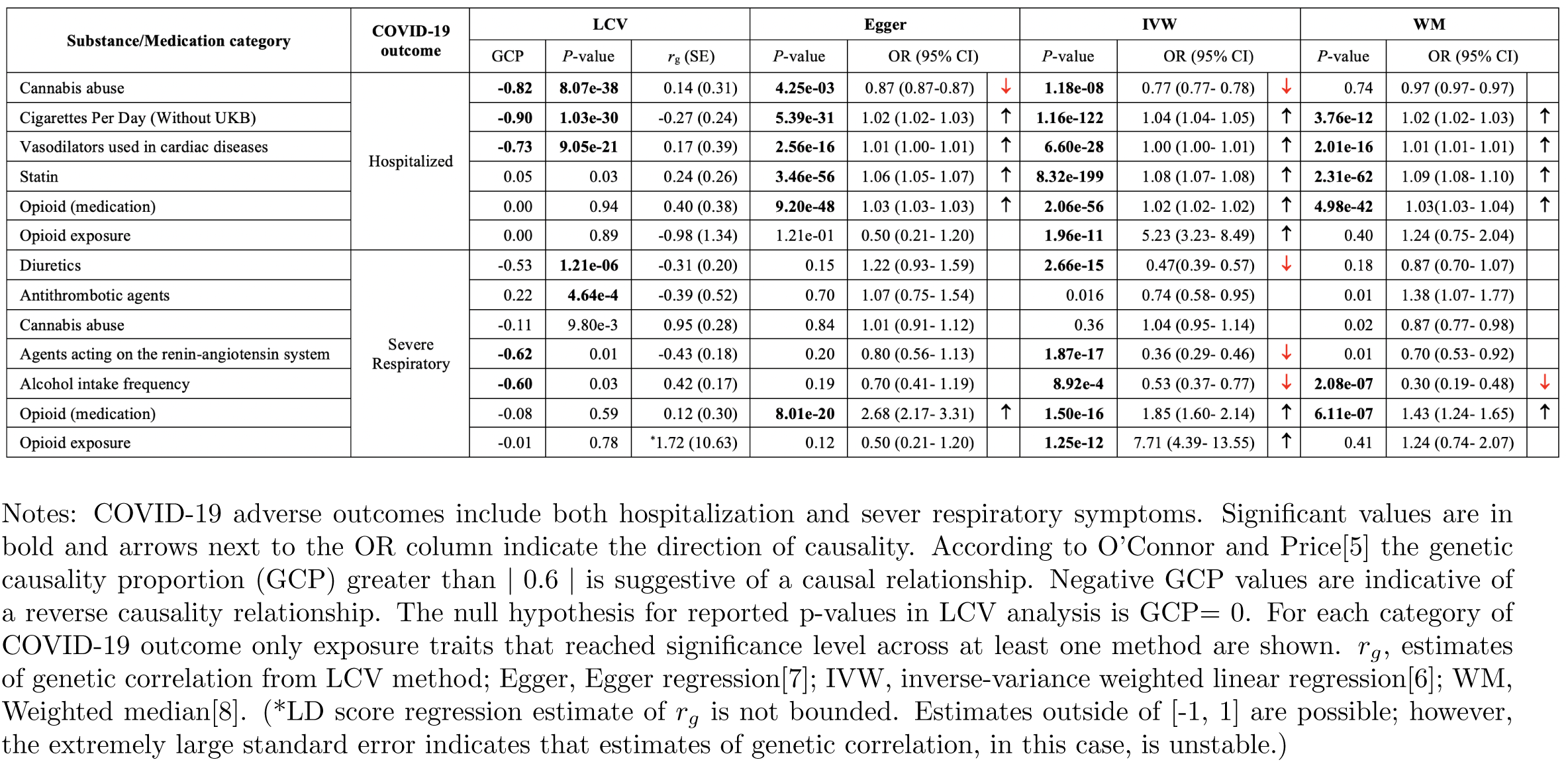
Summary of MR results for causal relationship between substance/medication traits (exposure) and COVID-19 adverse outcomes.

Consistent with the strong and significant impact of opioid use disorders (OUD) in *Wang et al*. analysis, we also identified a significant causality estimate for the opioid exposure trait (as a proxy for OUD) through the IVW regression method with a relatively similar effect size on both COVID-19 hospitalization and respiratory failure (Hospitalization: *OR*_*IVW*_ = 5.23,*P* _*IVW*_ = 1.96e-11; severe respiratory symptoms: *OR*_*IVW*_ = 7.71, *P*_*IVW*_ = 1.25e-12). Since IVW causality estimates are sensitive to horizontal pleiotropy, we further tested whether opioids as a medication (prescribed in the clinical setting) exert any causal effect on COVID-19 illness. To have a point of reference, we included medication classes that are primarily prescribed to treat high blood pressure and cardiovascular diseases (two well-known risk factors for severe COVID-1910). We observed a significant effect of opioid medications on both COVID-19 hospitalization and respiratory failure (**Table 1**). The comparison of causality odds ratios suggests that opioid medications have a strong effect on severe respiratory symptoms (*OR*_*Egger*_ = 2.68, *P*_*Egger*_ = 8.01e-20) while only minimally (up to ∼3%) increase the odds of hospitalization.

Our study showed that two classes of medications, including vasodilators and statin (used for cardiac diseases) are causally related to the risk of COVID-19 hospitalization (**Table 1**). The impact of vasodilators on the risk of hospitalization was almost negligible (up to 1% increase), but statin showed a stronger effect on the risk of hospitalization (*OR*_*Egger*_ = 1.06, *P*_*Egger*_ = 3.46e-56). The positive genetic correlation of both medication classes with COVID-19 hospitalization (vasodilators: *r*_*g*_ = 0.17, statin: *r*_*g*_ = 0.24) confirms the previously reported higher baseline prevalence cardiovascular conditions among hospitalized patients[8]. Neither of the two classes of medication showed a significant causality relationship with COVID-19 respiratory failure. We also detected a significant negative causal effect for diuretics and medication class acting on the renin-angiotensin system (RAAS) with the risk of COVID-19 reparatory failure. Since both medication classes are primarily prescribed for patients with hypertensive disease, this negative causal effect further underlines the importance of ACE inhibitors in controlling COVID-19 severe illness (diuretics: *OR*_*IVW*_ = 0.47, *P*_*I*_*V W* = 2.66e-15; RAAS: *OR*_*IVW*_ = 0.36, *P*_*IVW*_ = 1.87e-17).

Contrary to *Wang et al*., we detected evidence for a (negative) causal effect of cannabis abuse with both COVID-19 hospitalization and severe respiratory symptoms (**Table 1**). There is no clear mechanistic evidence linking cannabis use disorder (CUD) to COVID-19 symptoms, but we hypothesize that this negative causal effect is exerted through immunomodulatory pathways related to cannabinoid receptors. We found this observation interesting since *Wang et al*. also observed a protective association for lifetime CUD and COVID-19 (*OR* = 0.85, *P* = 0.006).

In conclusion, our MR analysis confirms the causal relationship between opioids and severe COVID-19 illness. However, our MR analysis questions the validity of the causal relationship between CUD and COVID-19 severe illness. Recently *Anil et al*.[9] showed treatment with cannabis compounds significantly reduces cytokine secretion in lung epithelial cells and, therefore, may be useful in alleviating severe symptoms among COVID-19 patients. The fact that a great deal of COVID-19 mortality is attributed to the disturbed immune regulation and cytokine storm, the possible modulatory impact of cannabinoids merits further investigation.

## Data Availability

All data produced are available online at:
https://www.covid19hg.org/results/r4/

https://github.com/RezaJF/COVID19_MR

## Data Availability

All data produced are available online at:
https://www.covid19hg.org/results/r4/

https://github.com/RezaJF/COVID19_MR

## Supplementary Note

To infer the causal relationship between SUD and COVID-19 adverse outcomes (Hospitalisation and severe respiratory symptom), we carried out a Mendelian Randomisation (MR) analysis. Unlike randomised trials that are costly, time prohibitive and, in the case of SUD, unethical to carry out, MR offers an amenable alternative for inferring causal relationships in a timely manner and efficiently. Furthermore, the application of MR in inferring causality also circumvents the biased conclusions occasionally associated with observational studies due to confounding and reverse causality.

The premise of MR relies on the association of exposure and outcome with genetic variants. Furthermore, MR can be carried out using only the GWAS summary statistics from the exposure and outcome traits. Since genetic variants are randomly segregated at conception, in MR analysis, genotypes are used as naturally occurring instruments. As such, a valid instrument is a variant associated with the exposure, but it is not associated with confounders of the exposure-outcome association. This instrumental variable is exclusively associated with the outcome via its effect on the exposure, revealing the causal relationship between the exposure and the outcome (**Figure S1**).

**Figure S1:**
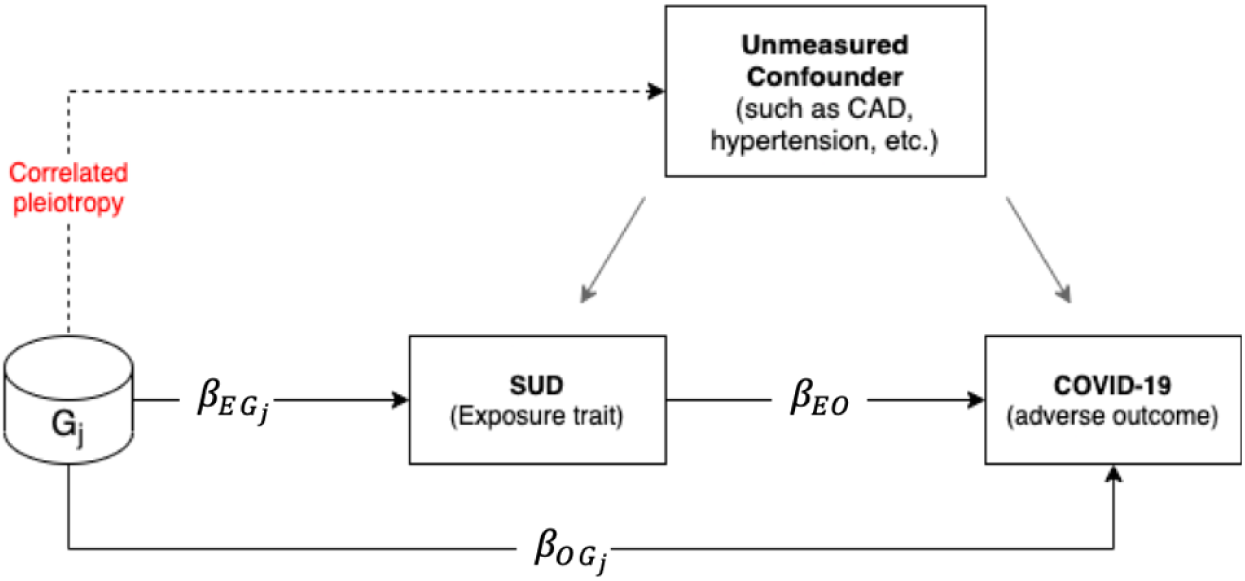
Schematic representation of assumptions underlying MR analysis.

The target of inference in MR analysis is *β*_*EO*_ (i.e. the causal effect of the exposure trait on the outcome). To circumvent the confounding effect of unmeasured traits (such as cardiovascular disorders and hypertension) that may be correlated with both the exposure and outcome (identified by grey arrows), we use genetic variants associated with both the exposure and outcome as the naturally randomized instruments. In this causal diagram, 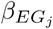 is the measure of association between the genetic variant *G*_*j*_ and the exposure trait (SUD in our analysis), 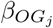 is the association of *G*_*j*_ with the outcome (i.e. COVID-19 hospitalization or severe respiratory symptom) and *β*_*EO*_ is the causal effect of exposure trait on the outcome. In our analysis, we make two important assumptions; 1) we assume that genetic variant *G*_*j*_ affects both the exposure and outcome through the same causal pathway, and 2) our randomized instrument *G*_*j*_ is not associated with any unmeasured confounder (i.e. there is no correlated pleiotropy).

### Mendelian Randomisation methods

In an MR analysis, in principle, we derive the causal effect (for each SNP) as the ratio of SNP-outcome association to SNP-exposure association. Specifically, the hypothesized relationship between the exposure and outcome for each instrumental variable *j* (out of *J*) is given by:

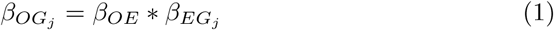

where 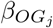 is the effect size of instrumental variable (*SNP*_*j*_) association with the outcome (in the case of our analysis covid-19 hospitalization or severe respiratory outcome), 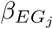 is the effect size of the association between the instrumental variable (*SNP*_*j*_) and the exposure (for example OUD or CUD in our analysis), and *β*_*OE*_ is the causal effect of **E** (exposure) on **O** (outcome). In GWAS summary statistics, all variants have been coded to reflect the positive SNP-exposure association; hence 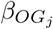 is always non-zero. Derivation of causal effect *β*_*OE*_ motivates the use of ratio so that for each instrumental variable *j*, the causal effect can be estimated by:

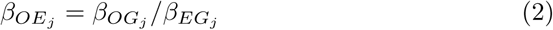

The WM (or IVW) method estimates the causal effect simply as a variance weighted (or an inverse variance weighted) average of all instrumental variable ratios 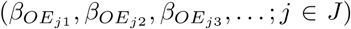. The Egger method extends this basic principle of MR to allow all variants to exert a pleiotropic effect on the outcome via the regression model based on:

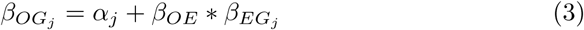

where *α*_*j*_ is the pleiotropic effect of instrumental variable SNP j.

The LCV method principally follows a similar framework as the Egger regression, but it is more mathematically involved and relies on the estimation of the Latent Causal Variable (LCV) that mediates the genetic correlation (*r*_*g*_) between the **E** and **O** traits. Under the LCV model, the relative proportions of heritability (between the exposure and outcome) that is attributable to the shared factor is calculated. To quantify the magnitude of heritability explained by the shared factor, O’Connor and Pric[5] introduce the “genetic causality proportion (GCP)” that ranges from -1 to 1 depending on the direction of causality.

Under this model, a fully genetically causal relationship (defined as 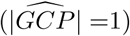 is inferred when the entire effect of the exposure trait on the outcome is derived from the genetic component underlying the shared factor 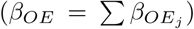. The LCV method computes the significance of GCP against the null hypothesis where 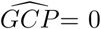. According to O’Connor and Pric[5], GCP estimates larger than 0.6 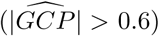 suggest a plausible causal relationship.

### Implementation of the MR analysis

We explored the causal relationship between the SUD and COVID-19 adverse outcome under four models, including LCV, Egger regression, inverse-variance weighted linear regression (IVW) and weighted median (WM).

We implemented our workflow in R. For investigating the causal relationship in LCV we used the R implementation of the model available at: https://github.com/lukejoconnor/LCV.

For the remaining models we used the TwoSampleMR package (v.5.6)[6] and MRPRESSO[10].

Codes underlying our analysis is available at: https://github.com/RezaJF/COVID19MR.

### Description of data source

We collected the summary statistics from published and ongoing GWAS studies across multiple resources. For each trait analysed in our study, a brief description of the data source is provided below:

1. **COVID-19 host genetics initiative (HGI):** We obtained GWAS summary statistics for “hospitalised COVID-19 cases vs. not hospitalised patients” and “very severe respiratory confirmed COVID-19 cases vs. population” from the meta-analysis round 4 of the COVID-19 host genetics initiative (https://www.covid19hg.org/about/). The COVID-19 host genetics initiative (HGI) is a collaborative project to identify genetic determinants of COVID-19 susceptibility, severity, and outcomes. Results from the GWAS analysis are available to download freely from the project website. The summary statistics for the hospitalisation record (B1_ALL) include meta-analysed GWAS weights across the 14,901,153 loci obtained from the analysis of 2,430 cases and 8,8478 controls with primarily European ancestry. The summary statistics for the severe respiratory symptom (A2 ALL) include meta-analysed GWAS weights across the 11,830,413 variants obtained from the analysis of 4,933 cases and 1,398,672 controls with primarily European ancestry.
2. **Psychiatric Genomics Consortium (PGC):** We obtained the GWAS summary statistics for opioid dependence, alcohol dependence and cannabis use disorder from the Psychiatric Genomics Consortium (https://www.med.unc.edu/pgc/download-results). These summary statistics are ascertained from the relevant studies carried out under the PGC auspice and are briefly described below:
  - **Opioid use disorder (OUD)** The GWAS summary statistics for “opioid dependence” include GWAS weights across 5,986,961 loci obtained from the analysis of 4,503 opioid-dependent, 4,173 opioid exposed and 32,500 opioid-unexposed controls. Details pertaining to the analysis and GWAS results are provided in the paper by Polimanti et al.[3].
  - **Alcohol use disorder (AUD)** We used the GWAS summary statistics for “alcohol use disorder” from Sanchez-Roige et al.[11]. The summary statistic includes meta-analyzed GWAS weights across 16,213,999 obtained from the analysis of 121,604 individuals in the UK biobank and 20,328 cases from the 23andMe. The authors used quantitative measures from the Alcohol Use Disorders Identification Test (AUDIT) to categorize participants as case and control.
  - **Cannabis use disorder (CUD)** The GWAS summary statistic for CUD includes meta-analysed GWAS weights across 11,535,788 variants from the analysis of 20,916 case samples and 363,116 controls from three cohorts, including the Psychiatric Genomics Consortium Substance Use Disorders working group, iPSYCH, and deCODE. For details related to the analysis, please see Johnson et al.[2].
3. **UK Biobank medication-use** We used GWAS summary-statistics across six categories of medication use from Wu et al.[4] (**Table S1**). These summary statistics are obtained from the analysis of self-reported medication use across 23 medication categories among the participants of the UK Biobank.

**Table S1.** Summary of medication-use GWAS.

| Data Source | Trait | No. of cases | No. of controls | No. of total tested GWAS variants | No. independent GWAS hit ($P \leq 5E-8$) | Reference |
| --- | --- | --- | --- | --- | --- | --- |
| HGI | Hospitalised COVID-19 cases vs. not hospitalised | 2,430 | 8,8478 | 14,901,153 | 14 | www.covid19hg.org |
|  | Very severe respiratory confirmed COVID-19 cases vs. population | 4,933 | 1,398,672 | 11,830,413 | 542 |  |
| PGC | Opioid use disorder (OUD) | 8,676 | 32,500 | 5,986,961 | 20* | Polimanti *et al.* 2020 |
|  | Alcohol use disorder (AUD)* | 121,604 UK Biobank<br>20,328 23&Me |  | 16,213,999 | 83 | Sanchez-Roige *et al.* 2019 |
|  | Cannabis use disorder (CUD) | 20,916 | 363,116 | 11,535,788 | 752 |  |
| UK Biobank | Agents acting on the renin-angiotensin system | 62,752 | 174,778 | 7,288,504 | 184 | Wu *et al.* 2019 |
|  | Antithrombotic agents | 67,653 | 85,986 |  | 13 |  |
|  | Diuretics | 34453 | 194633 |  | 102 |  |
|  | Opioid (medication) | 22982 | 55826 |  | 3 |  |
|  | Statin | 73475 | 216910 |  | 97 |  |
|  | Vasodilators used in cardiac diseases | 5546 | 237113 |  | 3 |  |

♣ In Sanchez-Roige *et al.*, AUD is treated as a quantitative trait. The authors calculated the Alcohol Use Disorder Identification Test (AUDIT) score across all participants and used different thresholds to dichotomise the cohort into alcohol consumption (AUDIT-C, $N = 121,604$ ) and alcohol problem (AUDIT-P, $N = 121,604$ ) sub-cohorts.
♠ Given the comparatively lower number of total variants tested for OUD, we used the lenient $p$ -value threshold $p = 5E-6$ to select instrumental variables.

